# Occupational Exposure to Heat and Wildfire Smoke in California Correctional Facilities

**DOI:** 10.1101/2025.08.04.25332973

**Authors:** Rachel Sklar, Samantha Kramer, Fred Lurmann, Marissa Childs, Stefano Bertozzi, Elizabeth Noth

## Abstract

1.

**Objectives:** To assess long-term trends in outdoor heat and wildfire smoke exposure at 410 correctional facilities in California from 2000–2023.

**Methods:** We used ERA5-Land data to calculate daily wet-bulb globe temperature (WBGT) and identified days exceeding the NIOSH 28°C threshold. Wildfire smoke exposure was estimated using NOAA’s Hazard Mapping System (HMS). Temporal trends in daily heat and smoke exposure were modeled using linear regression for each facility.

**Results:** Smoke-affected days rose statewide from 4.9% (2006–2010) to 12.5% (2019– 2023), with Northern California experiencing the highest burden and steepest increases. Days above 28°C WBGT increased from 27.3% to 29.8% statewide, with the Central Valley and desert regions showing the highest current burden and coastal areas experiencing the steepest increases. Coastal facilities show emerging heat risks, and some regions—especially the Central Valley and Sierra foothills—face dual exposure to both hazards.

**Conclusions:** Correctional officers and incarcerated workers face rising environmental risks, with regional variation in exposure. Facilities with concurrent heat and smoke burdens must balance cooling and filtration needs. Resilience planning and occupational protections must evolve to meet facility- and region-specific challenges.

## 2. Introduction

Correctional facilities in California are often located in regions increasingly affected by extreme heat and wildfire smoke. ^1,2^ As of 2023, over 33,740 correctional officers and 64,788 incarcerated workers were employed within these settings in California. ^3,4^ Many correctional facilities were not built to withstand modern climate stressors. Their aging, heat-retaining concrete structures and outdated HVAC systems offer limited protection. ^5,6^ These environmental hazards are compounded by demanding job requirements, making it difficult to mitigate risks for staff and incarcerated individuals alike.

Existing studies have begun to examine heat exposure in U.S. prisons, though the literature remains sparse.^2,7^ Research on wildfire smoke exposure in correctional settings remains especially limited. This study explores current and long-term patterns in both heat and wildfire smoke across California correctional facilities, highlighting the need for targeted research on occupational exposures and controls. We focus on working populations given limited occupational research in correctional settings, though incarcerated residents likely face greater exposure due to living continuously in these facilities.

## 3. Methods

Correctional facilities were identified using North American Industry Classification System (NAICS) codes. Using ArcGIS, we mapped the locations of all 410 identified California correctional facilities spanning federal, state, county, local, and multi-jurisdictional institutions. Facilities included adult and juvenile detention centers with varying security classifications. Heat exposure data were obtained from the ERA5-Land reanalysis dataset spanning 2006-2023, with daily maximum wet-bulb globe temperature (WBGT) values calculated using temperature, humidity, and solar radiation data at each facility location.^8^ For each facility and year, we calculated the percentage of days exceeding the National Institute for Occupational Safety and Health (NIOSH) recommended threshold of 28°C WBGT for outdoor work activities. To assess long-term trends in heat exposure, we fitted linear regression models for each facility, using percentage of hot days as the dependent variable and year as the independent variable across the full 24-year study period (2000-2023). We calculated the total change in heat exposure over the study period by multiplying the annual rate of change (slope coefficient) by 23 years.^2^ For smoke, we calculated the percentage of days each year with detectable smoke from 2006–2023 using NOAA’s Hazard Mapping System (HMS) plumes and applied the same linear regression approach to assess the trend in smoke exposure over a 17-year period. ^9^

## 4. Results

On average, the percentage of smoke-affected days rose from 4.9% (2006–2010) to 12.5% (2019–2023), reflecting a marked increase in smoke exposure in the state. The steepest increases and highest current smoke days are concentrated in Northern California, particularly the northern Sierra foothills and adjacent valleys (Figure 1a). Statewide, the average percentage of heat days above the 28°C WBGT threshold rose from 27.3% (2006–2010) to 29.8% (2019–2023), reflecting a more modest increase in outdoor heat exposure. The steepest heat increases appear in coastal Southern California and the LA basin, while the Central Valley and desert regions currently experience the highest current heat levels (Figure 1b). Several coastal Southern California facilities show steep upward trends in heat exposure despite relatively low current heat levels, indicating emerging thermal risk. Some regions have mismatched burdens such as the northern part of the state with high smoke but moderate heat, and inland southern areas with high heat but moderate smoke. The Central Valley and Sierra foothills face a dual burden, with high and rising levels of both (Figure 1).

**Figure 1.**
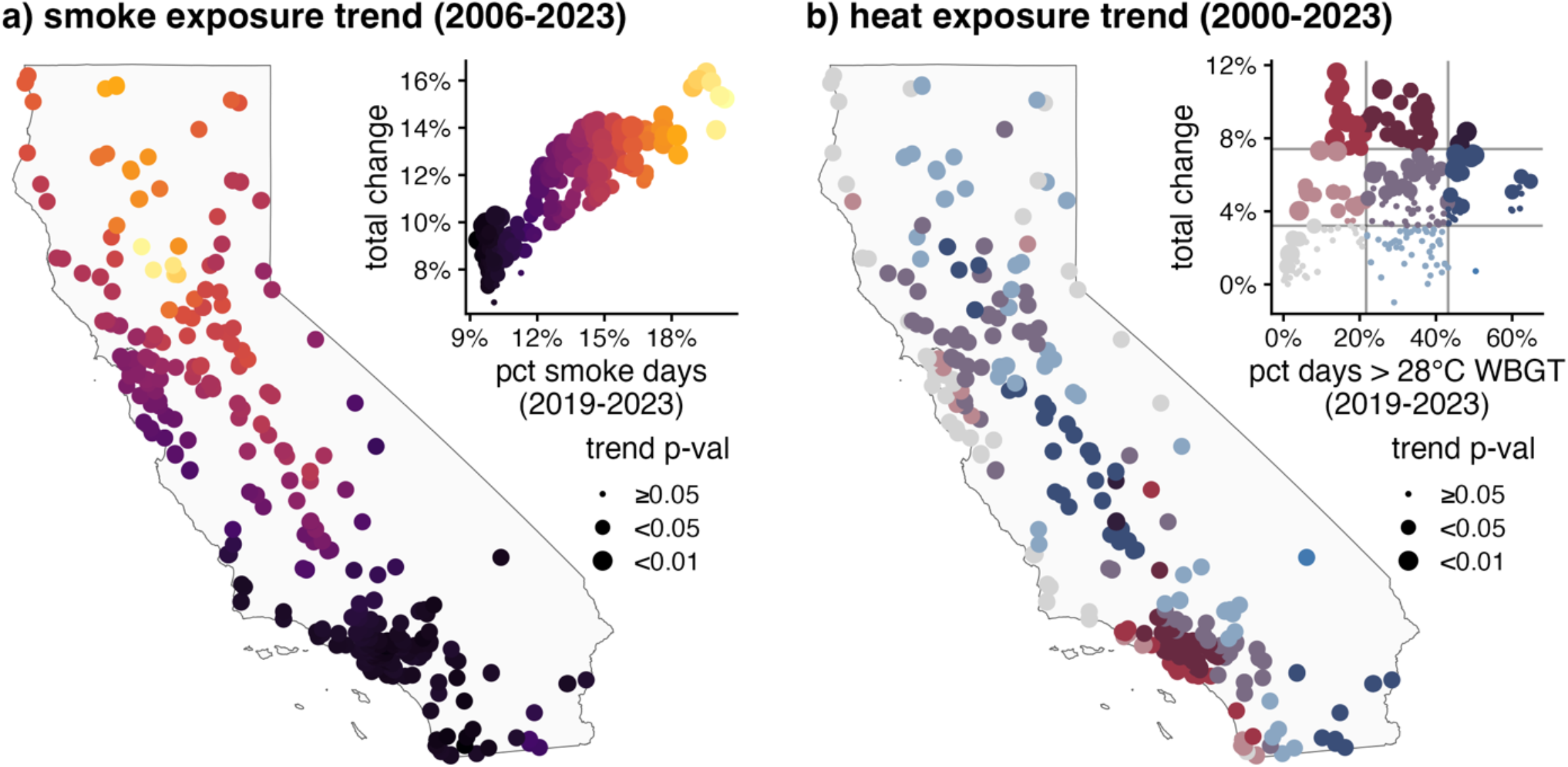
Environmental exposure trends at California correctional facilities. (a) Smoke exposure trends (2006-2023) showing current smoke levels and magnitude of 17-year change across facilities. (b) Heat exposure trends (2000-2023) showing current heat levels and magnitude of 23-year change. The 410 correctional facilities shown span federal, state, county, local, and multi-jurisdictional institutions. Current heat and smoke levels are calculated as an annual average over the last 5 years in the sample (2019-2023).

Notably, the relationship between current and long-term exposure differs by hazard. For smoke, high current exposure correlates strongly with the upward trend over the years. However, for heat, the greatest upward trend is seen in places with low to moderate current burden. Locations with already high burden may also see increases in the intensity of heat days not captured in the metric of days over °C WBGT.

## 5. Discussion

People who live and work in California correctional facilities are facing growing health risks from heat and wildfire smoke exposure. These risks may be magnified by aging and inadequate infrastructure.^5,6^ Many occupational factors—such as personal protective equipment (PPE) use, limited environmental control, and extended shifts—may also elevate risk of heat and smoke exposure for both correctional officers and incarcerated workers. Correctional officers wear heavy protective equipment that inhibits heat dissipation.^10^ They may work overtime shifts due to staffing shortages which may increase heat exposure especially during emergency incidents. ^1^

During wildfire events, officers and incarcerated workers may face a tradeoff between wearing respiratory protection, which can worsen heat stress, or forgoing masks and risking smoke exposure.^11^ Underlying health conditions and psychotropic medication use further elevate heat-related illness risk among incarcerated workers.^2^

Regional variability in smoke and heat trends suggests that Southern California facilities may need to prioritize extreme heat mitigation, while Northern California institutions may face greater challenges from increasing wildfire smoke, even in the absence of significant heat trends. A small but important subset of facilities, primarily in the Central Valley and Sierra foothills, faces concurrent increases in both heat and smoke exposure raising concerns about the health effects of co-exposure and the challenge of balancing cooling needs with air filtration when outdoor air intake may worsen indoor air quality. These differences highlight the need for facility-specific risk assessments and tailored mitigation strategies that account for the distinct environmental hazards facing each region. Additionally, emerging heat burdens in parts of coastal California, areas historically considered low risk, raise questions about whether the facilities in these regions are adequately equipped with the infrastructure and plans needed to mitigate these exposures.

This study did not directly assess indoor environmental conditions where both free and incarcerated workers may spend most of their time at work. Future work will examine how these outdoor estimates translate into occupational risks indoors. Additionally, while the HMS smoke plume data provide consistent, long-term spatial coverage, they are limited in vertical resolution and may not accurately capture ground-level concentrations, but are similar to patterns in estimated ground-level wildfire smoke fine particulate matter.^12^ As a result, our estimates may over- or underestimate smoke days relevant for exposure assessment. Future analyses will incorporate additional data sources to better characterize surface-level smoke conditions across correctional settings.

## 6. Public Health Implications

Environmental hazards like extreme heat and wildfire smoke pose serious threats to the health, safety, and productivity of both correctional officers and incarcerated workers. These conditions can lead to more injuries and illnesses, make it harder to perform physical or decision-intensive tasks, and strain operations and security. For correctional officers, this may mean increased medical leave, staffing shortages, and financial pressures on agencies to respond to workers compensation claims. For incarcerated workers, exposure to heat and smoke may limit participation in job assignments and programming, potentially reducing opportunities for skill-building and rehabilitation. Addressing these risks through targeted mitigation strategies is essential to ensure safer, more resilient correctional environments for all. These risks cannot be addressed through one-size-fits-all solutions, as the correctional facilities explored here differ in size, infrastructure type, and capacity to implement exposure controls.

## Data Availability

Select data produced in the present study are available upon reasonable request to the authors.

## References

1. LeMasters K, Haber LA. The hidden crisis of incarcerated individuals during wildfires. The Lancet Regional Health - Americas. 2025;43:101032. doi:10.1016/j.lana.2025.101032

2. Tuholske C, Lynch VD, Spriggs R, et al. Hazardous heat exposure among incarcerated people in the United States. Nat Sustain. 2024;7(4):394–398. doi:10.1038/s41893-024-01293-y

3. Occupational Employment and Wages, May 2023: 33-3012 Correctional Officers and Jailers. U.S. Bureau of Labor Statistics. Accessed May 28, 2025. https://www.bls.gov/oes/2023/may/oes333012.htm#(1)

4. Macomber J, Singh D, Aghakhanian DA, Davison D. California Prison Industry Board. California Prison Industry Authority

5. Jay O, Capon A, Berry P, et al. Reducing the health effects of hot weather and heat extremes: from personal cooling strategies to green cities. The Lancet. 2021;398(10301):709–724. doi:10.1016/S0140-6736(21)01209-5

6. Sklar R, Noth E, Kwan A, Sear D, Bertozzi S. Ventilation conditions during COVID-19 outbreaks in six California state carceral institutions. PLOS ONE. 2023;18(11):e0293533. doi:10.1371/journal.pone.0293533

7. Ovienmhada U, Hines M, Krisch M, Diongue AT, Minchew B, Wood DR. Spatiotemporal Facility-Level Patterns of Summer Heat Exposure, Vulnerability, and Risk in United States Prison Landscapes. GeoHealth. 2024;8(9):e2024GH001108. doi:10.1029/2024GH001108

8. Spangler KR, Liang S, Wellenius GA. Wet-Bulb Globe Temperature, Universal Thermal Climate Index, and Other Heat Metrics for US Counties, 2000–2020. Sci Data. 2022;9(1). doi:10.1038/s41597-022-01405-3

9. Vargo JA. Time Series of Potential US Wildland Fire Smoke Exposures. Front Public Health. 2020;8. doi:10.3389/fpubh.2020.00126

10. Hudson S, Ridland L, Blackburn J, Monchuk L, Ousey K. The comfort and functional performance of personal protective equipment for police officers: a systematic scoping review. Ergonomics. 2024;67(10):1317–1337. doi:10.1080/00140139.2024.2302957

11. Li Y, Tokura H, Guo YP, et al. Effects of wearing N95 and surgical facemasks on heart rate, thermal stress and subjective sensations. Int Arch Occup Environ Health. 2005;78(6):501–509. doi:10.1007/s00420-004-0584-4

12. Childs ML, Li J, Wen J, et al. Daily Local-Level Estimates of Ambient Wildfire Smoke PM2.5 for the Contiguous US. Environ Sci Technol. 2022;56(19):13607–13621. doi:10.1021/acs.est.2c02934

